# Ocular Surface Microbiome: Influences of Physiological, Environmental, and Lifestyle Factors

**DOI:** 10.1101/2024.07.01.24309728

**Authors:** Vincenzo Rizzuto, Marzia Settino, Giacomo Stroffolini, Giuseppe Covello, Juris Vanags, Marta Naccarato, Roberto Montanari, Carlos Rocha de Lossada, Cosimo Mazzotta, Carlo Adornetto, Miguel Rechichi, Francesco Ricca, Gianluigi Greco, Guna Laganovska, Davide Borroni

**Affiliations:** Department of Ophthalmology, Riga Stradins University, Riga, Latvia; Eyemetagenomics Ltd., London, United Kingdom; Centro Oculistico Borroni, Gallarate, Italy; Clinic of Ophthalmology, P. Stradins Clinical University Hospital, Riga, Latvia; School of Advanced Studies, Center for Neuroscience, University of Camerino, Camerino, Italy; Latvian American Eye Center (LAAC), Riga, Latvia; Iris Medical Center, Cosenza, Italy; Department of Infectious-Tropical Diseases and Microbiology, IRCCS Sacro Cuore Don Calabria Hospital, Verona, Italy; Department of Surgical, Medical, Molecular Pathology and Critical Care Medicine, University of Pisa, Pisa, Italy; Ophthalmology Department, QVision, Almeria, Spain; Ophthalmology Department, Hospital Regional Universitario of Malaga, Malaga, Spain; Department of Surgery, Ophthalmology Area, University of Seville, Seville, Spain; Siena Crosslinking Center, Siena, Italy; Departmental Ophthalmology Unit, USL Toscana Sud Est, Siena, Italy; Postgraduate Ophthalmology School, University of Siena, Siena, Italy; Centro Polispecialistico Mediterraneo, Sellia Marina, Italy; Department of Mathematics and Computer Science, University of Calabria, Rende, Italy; Pharmacology Institute, Heidelberg University Hospital, Heidelberg, Germany

**Keywords:** Microbiome, Bioinformatics, Confounding Factors, Ocular Microbial Composition, Microbial composition, 16S rRNA gene profiling

## Abstract

**Purpose:** Purpose: The ocular surface (OS) microbiome is influenced by various factors and impacts ocular health. Understanding its composition and dynamics is crucial for developing targeted interventions for ocular diseases. This study aims to identify host variables, including physiological, environmental, and lifestyle (PEL) factors, that influence the ocular microbiome composition and establish valid associations between the ocular microbiome and health outcomes.

**Methods:** The 16S rRNA gene sequencing was performed on OS samples collected using eSwab. DNA was extracted, libraries prepared, and PCR products purified and analyzed. PEL confounding factors were identified, and a cross-validation strategy using various bioinformatics methods including Machine learning was used to identify features that classify microbial profiles.

**Results:** Nationality, sport practice, and eyeglasses usage are significant PEL confounding factors influencing the eye microbiome. Alpha-diversity analysis showed higher microbial richness in Spanish subjects compared to Italian subjects and higher biodiversity in sports practitioners. Beta-diversity analysis indicated significant differences in microbial community composition based on nationality, age, sport, and eyeglasses usage. Differential abundance analysis identified several microbial genera associated with these PEL factors. ML approach confirmed the significance of nationality in classifying microbial profiles.

**Conclusion:** This study underscores the importance of considering PEL factors when studying the ocular microbiome. Our findings highlight the complex interplay between environmental, lifestyle, and demographic factors in shaping the OS microbiome. Future research should further explore these interactions to develop personalized approaches for managing ocular health.

**Key Points:**

- Identify confounding factors influencing the ocular microbiome composition;
- Characterize the ocular surface microbiome;
- Analyse 16S rRNA gene sequencing data from ocular surface samples;
- Perform Diversity Analysis (i.e.; Alpha-diversity and Beta-diversity) and Difference Abundance Analysis;

## Introduction

More than ten years have elapsed since the inception of the Human Microbiome Project [1], aiming to establish a baseline genome of the normal human microbiota across five principal regions—oral cavity, airways, skin, gastrointestinal tract, and vagina [2]. Specific human habitats delineate unique microbial niches. Each microbial community is shaped not only by host-related elements like pH and oxygen levels but also by external influences such as diet and exposure to antibiotics [3]. As sequencing technologies advance, large scale microbiome studies are rapidly increasing. Although “microbiota” and “microbiome” are frequently used as synonyms, there are differences between the two terms. The microbiota is the wide variety of microorganisms living in a specific environment such as oral, ocular and intestinal. Microbiome encompasses the collection of genomes of the entire habitat of microorganisms in the environment.

Therefore, with respect to the microbiota, microbiome includes a wider spectrum. Metagenomics is a research discipline based on the development of next-generation sequencing (NGS) technology that is able to detect the genetic material (i.e.; microbiome) of several microorganisms recovered from biological samples [4]. Microbiome profiling is commonly conducted using the reads of specific marker genes such as the evolutionary conserved 16S rRNA gene [5]. Microbiome profiling is revolutionizing our understanding of biological mechanism underlying the onset and development of several diseases. Several evidences in the biomedical literature suggest that microbiome composition is influenced by the subject physiological, environmental and lifestyle (PEL) confounding factors such as the nationality, practice of sport, use of eyeglasses, smoking addiction, etc. A metagenomics study reveals that in healthy centenarians there are differences between sexes by comparing the metagenomic characterizations of the gut microbiome [6]. Smoking has been shown to alter the gut microbiome composition and contribute to the overall adverse health impact [7].

The gut microbiome has been extensively investigated and it is actually considered one of the key elements contributing to the human health [8]. Otherwise, the human eye microbiome has been only partially and insufficiently explored [9, 4, 10]. The human ocular surface is colonized by a diverse community of microorganisms, collectively known as the ocular surface microbiome. This microbiome varies between individuals and is influenced by host factors such as age, diet, hygiene, and geographic location [11, 12]. Recent studies have indicated that changes in the ocular surface microbiome can be associated with various ocular diseases, including conjunctivitis, dry eye, keratoconus and microbial keratitis [13,14,15,16,17,18]. The Ocular Microbiome Project, initiated in 2010, aimed to investigate the relationship between the ocular microbiome and ocular diseases [19].

Metagenomic studies have revealed that the healthy ocular surface (OS) typically exhibits a relatively stable microbiota characterized by low diversity. A “core” microbiota, consisting of a few taxa, is shared among all individuals [20], encompassing commensal, environmental, and potentially pathogenic bacteria [21]. The predominant phyla observed on the OS include Proteobacteria, Actinobacteria, and Firmicutes [11,12,22,23,24,25,26,27,28]. At the genus level, commonly found taxa comprise Pseudomonas, Propionibacterium, Bradyrhizobium, Corynebacterium, Acinetobacter, Brevundimonas, Staphylococci, Aquabacterium, Sphingomonas, Streptococcus, Streptophyta, and Methylobacterium. The composition of the microbiota residing on the healthy ocular surface changes with several factors, and there is no consensus on whether a core microbiome exists [29, 30, 31].

Yet, while the complete characterization of the ocular microbiome in healthy individuals remains incomplete, our prior study aimed to categorize the diverse bacterial community profiles existing harmoniously within a healthy eye [32]. We introduced the term “eye community state type” (ECST) to describe these varied profiles.

While the influence of the microbiome on human health has become increasingly clear, various challenges, some of which resemble those encountered in other high-throughput studies, have yet to be fully addressed. A confounder factor is a variable that influences both the dependent variable and independent variable causing a biased causal association [33]. Confounding factors such as physiological (e.g.; sex, age), environmental (e.g; nationality) and lifestyle (e.g.; smoking) could create spurious correlation between microbiome composition and diseases and thus they may bias the results by obscuring the differential features in the case-control study [34,35,36,37,38,39].

Machine learning (ML) are becoming a standard tool for identifying disease biomarkers. A biomarkers is a measurable feature that can be objectively measured and evaluated as an indicator of some biological state or condition, such a disease status. Biomarkers play an important role in precision medicine, for instance, by predicting the disease-related patient outcome [40].

In spite of these promising benefit, machine learning models could be driven by confounding factors and thus also to capture single or multiple confounding effect correlated with the outcome instead of capturing only the features specific to the outcome (i.e.; biomarkers) [41]. Therefore, confounding factors should be taken into consideration especially for ML-based approaches to perform causal inference for establishing a valid causal associations between the ocular microbiome composition and health outcomes as well as developing benchmark values [42].

Accounting for confounding in the microbiome is particularly difficult due to the high number of variables that can potentially influence the microbiome composition. Differential abundance analysis (DAA) is the most widely used approach to identify differences in the microbiome composition between samples groups [43]. In general, the most commonly methods to perform DAA fall into four main categories: a) simple statistical tests; b) RNA-seq based methods; c) metagenomic based methods and d) Supervised machine learning methods.

RNA-seq based methods (e.g.; DESeq) are adapted from RNA sequencing (RNA-seq) studies and they are widely used in the microbiome context because they benefit from well-developed software packages, wide acceptance, and a robust theoretical basis [44, 45]. However, specific features of the microbiome data make the RNA-seq based methods difficult to apply in complex study designs. Indeed, compared to RNA-seq data, metagenomic data are characterized by high dimensionality, uneven sequencing depth and data sparsity [46]. Typically, microbiome datasets contain a very large amount of complex, sparse and heterogeneous data (i.e.; microbial taxa) and a small number of samples are collected. Therefore RNA-seq based methods should be used with caution in the microbiome context [47, 48, 49].

Metagenomic based methods (e.g.; LEfSe) have been developed specifically for dealing with microbiome data. These methods include several sensitive tests that have greater power to detect differential features than simple statistical tests. Supervised learning (SL) methods can be applied effectively to microbiota for classification tasks [50]. SL involves training a ML model on a labeled dataset (i.e.; each data point has a corresponding label or output value) so that the model learns to predict the output for new input data. Random forest (RF), logistic regression (LR) and support vector machine (SVM) are SL algorithms that can be used to predict the belonging group for each sample (i.e.; classify samples) based on its metagenomic profile [51]. Prediction performance evaluation metrics such as Accuracy, the Area Under Receiver Operating Characteristic curve (ROC-AUC) and the Error Rate (i.e., the proportion of items that are incorrectly classified by the ML model) are used to evaluate the performance of the final classifier model. All of these methods have their own advantages and disadvantages, and to date there is no a gold standard [52].

This article aims to provide a substantial contribution to understanding the causal association between PEL confounding factors and microbiome composition to characterize the healthy eye microbiome. To obtain a good reliability level of the metagenomic data analysis results, we use a cross-validation strategy based on different bioinformatics methods to explore how the microbial communities partition with PEL confounding factors.

The rest of the paper is organized as follows. Section 2 describes the analysis carried out, the data collections and the subjects recruitment, Section 3 presents our findings, Section 4 compares the DAA results, Section 5 discusses the findings and Section 6 concludes the article.

## Materials and Methods

Bioinformatics Analysis is performed through the use of several R packages. Phyloseq (version 1.42) is used for representing and preprocessing microbiome data. Data are saved as a phyloseq object consisting of 2668 taxa and 135 samples. Diversity analysis (i.e.; Alpha and Beta diversity statistical tests) is performed by Stats package (version 4.2.2) that provides the kruskal.test and pairwise.wilcox.test functions. Furthermore, Vegan package (version 2.6) that implements the permutational multivariate analysis of variance (PERMANOVA) by the “adonis2” function [53] is used for diversity analysis. Difference Abundance analysis is performed by DESeq2 (version 1.38.3) and microbiomeMarker (version 1.4) packages [54, 55].The ocular surface microbiome composition is assessed with respect to the eleven probable PEL confounding factors reported in the Tables 1,2, They are categorical variables that can assume binary (e.g.; Smoking) or multi-class values (e.g.; BMI).

**Table 1.** PEL Factors, their Categories, and Values. Y stands for “Yes”; N stands for “No”; ES stands for Spanish; ITA stands for Italian.

| PEL Confounding Factors | Categories | Values |
| --- | --- | --- |
| Sex | Male (M), Female (F) | Male/Female |
| Eyeglasses | Yes (Y): Wears eyeglasses<br>No (N): Does not wear eyeglasses | Y/N |
| Smoking | Yes (Y): Smoker<br>No (N): Non-smoker | Y/N |
| Computer Usage | Yes (Y): Spends more than 5 hours daily on a computer<br>No (N): Spends less than 5 hours daily on a computer | Y/N |
| Allergy | Yes (Y): Has allergies<br>No (N): Does not have allergies | Y/N |
| Sport | Yes (Y): Engages in physical activity more than 3 times a week<br>No (N): Engages in physical activity less than 3 times a week | Y/N |
| Hypertension | Yes (Y): Diagnosed with hypertension<br>No (N): Not diagnosed with hypertension | Y/N |
| Reflux | Yes (Y): Suffers from gastroesophageal reflux (self-reported)<br>No (N): Does not suffer from gastroesophageal reflux | Y/N |
| Nationality | ES: Spanish nationality<br>ITA: Italian nationality | ES/ITA |

**Table 2.** Table reports the values that BMI can assume. The third column reports the range of BMI values.

| PEL confounding factors | Values | Range |
| --- | --- | --- |
| BMI | Underweight | $0 < \text{bmi} < 18$ |
| | Normal weight | $18 \leq \text{bmi} < 24$ |
| | Overweight | $24 \leq \text{bmi} < 29$ |
| | Obese | $\text{bmi} \geq 29$ |

### Generation of PEL Classes

The PEL variables values were collected using selfreported questionnaires during participant recruitment. The physiological, environmental, and lifestyle (PEL) factors are defined in the Tables 1,2,3.

**Table 3.**
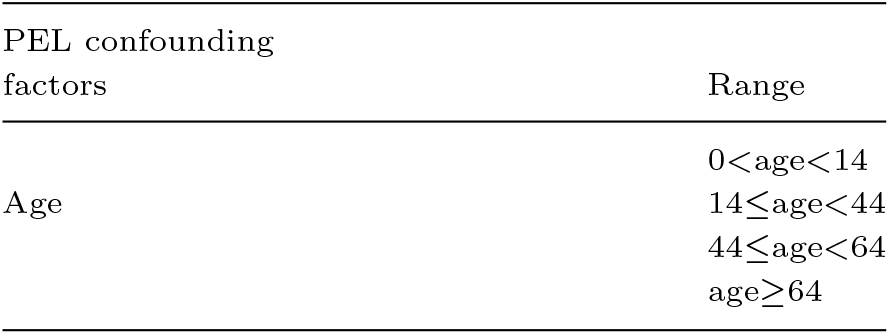
Table reports the values that Age can assume. Note that unlike BMI, age values coincide with the age range.

| PEL confounding factors | Range |
| --- | --- |
| Age | $0 < \text{age} < 14$ |
| | $14 \leq \text{age} < 44$ |
| | $44 \leq \text{age} < 64$ |
| | $\text{age} \geq 64$ |

### Data Collection and Subject Recruitment

A total of 135 subjects were recruited for this study. Participants were enrolled from several ophthalmology clinics, including Virgen de las Nieves University Hospital (Granada, Spain), Emilia-Romagna Eye Bank (Bologna, Italy), and Centro Polispecialistico Mediterraneo (Sellia Marina, Italy)[4]. Approval for this study is granted by the institutional review board of Riga Stradins University (nr.29/20092016). Participant recruitment occurred at several ophthalmology clinics: Virgen de las Nieves University Hospital (Granada, Spain), Emilia-Romagna Eye Bank (Bologna, Italy), and Centro Polispecialistico Mediterraneo (Sellia Marina, Italy). The study adhered to the principles outlined in the Declaration of Helsinki and received informed consent from all human subjects involved. Healthy individuals above 18 years old were included based on specific criteria and Ocular Surface Disease Index (OSDI) scores.

Healthy subjects were defined as individuals without a history of ocular surgery, allergies, ocular inflammation, contact lens use, ocular surface diseases, meibomian gland dysfunction, recent antibiotic intake within 6 months, systemic medication (as it might affect ocular microbiota), having a BMI index between 18.5 and 24.9, and normal blood glucose levels [56]. Eligible candidates meeting these criteria were invited to participate, and informed consent was obtained from each participant at the study’s outset.

### Sampling Technique, DNA Extraction, PCR Amplification, Library Preparation, and Amplicon Sequencing

Samples were collected using an eSwab containing 1 mL of Liquid Amies Medium (Copan Brescia, Italy). The eSwab was gently applied to the inferior eye surface, moving twice from “limbus to fornix to limbus.” To prevent any impact on the eye microbiota, no fluorescein or anesthetics were used [57] [58]. DNA extraction was performed using the QIAamp DNA Microbiome Kit (QIAGEN, Hilden, Germany) following the manufacturer’s protocols. Quantification of extracted DNA was conducted with the Agilent TapeStation 4150 utilizing Genomic DNA ScreenTape (Agilent Technologies, Santa Clara, CA, USA). Library preparation involved the Ion 16S Metagenomics Kit (Thermo Fisher, Waltham, MA, USA). Briefly, bacterial genomic DNA was diluted to 2 ng/mL, with 2 mL used for library preparation. Amplification was achieved through two primer pools targeting the V2-4-8 and V3-6, 7–9 regions of 16S rDNA. Post-amplification, PCR products underwent purification, end-repair, and barcode ligation. The final step included library amplification and pooling to achieve a concentration of 26 pM. Template preparation was executed with Ion Chef following the Ion 510, Ion 520, and Ion 530 Kit-Chef protocol.

Sequencing of the amplicon libraries was conducted using the Ion Torrent S5 system (Thermo Fisher, Waltham, MA, USA) on a 520 or 530 chip, following the supplier’s guidelines. Post-sequencing, individual sequence reads underwent filtering using Ion software to eliminate low-quality and polyclonal sequences. Sequences matching the IonXpress adaptor were automatically trimmed. All S5 quality-approved, trimmed, and filtered data were exported as bam files.

### Data Processing

Data processing is the process of converting raw data into a useful format for further analysis. Operational taxonomic unit (OTUs) is used to represent the Microbial count from 16S rRNA sequencing and it is considered the basic unit used in the numerical taxonomy, i.e.; OTUs define “mathematically” the taxa [59, 60].

To improve the downstream statistical analysis by eliminating artifactual bias in the original measurements [61], data processing is performed to remove low quality or uninformative OTUs which are likely due to sequencing errors or low-level contaminations in PCR amplicon [62]. In particular, low-abundance OTUs (i.e.; OTUs that are not present in at least 20% of the samples) and sparse OTUs (i.e.; OTUs with *>* 90% of zeros) are removed [63]. Furthermore, median sequencing depth is used to normalize the number of reads in each sample. Microbial abundance data are normalized by the Counts Per Million (CPM) method before performing the Difference Abundance analysis [64].

### Diversity Analysis

Microbial diversity refers to within-sample and between-sample diversity, named respectively Alpha-diversity and Beta-diversity [65, 66].

**Alpha-diversity** takes into account the number of different taxa that are found in each sample. We estimated Alpha-diversity using two metrics: Observed OTUs and Shannon index. The first is the most simple alphadiversity metric. It just counts up the number of different taxa observed in a sample at a given taxonomic level. The latter (a.k.a. Shannon’s diversity index and Shannon entropy) is an estimator for both taxa richness and evenness, but with weight on the richness. In addition, statistical analysis on alpha-diversity is performed by the Kruskal–Wallis test (for multi-class comparison) and Pairwise Wilcoxon Rank Sum Tests with Bonferroni correction (for pairwise comparison).

**Beta-diversity** measures the inter-individual diversity (i.e.; difference in microbial composition between samples) [67]. We explore Beta-diversity by the Principal Coordinates Analysis (PCoA) based on Bray-Curtis dissimilarities. PCoA is a non-linear dimension reduction technique based on euclidean distances between samples and it is a generalized version of the Principal Component Analysis (PCA).

### Difference Abundance Analysis

To identify the microbial taxa whose abundance covaries with the PEL confounding factors, the Difference Abundance Analysis is performed through a multi-approach based strategy. Differences in microbial abundance are explored through the following four methods:

- **Statistical (non-parametric) tests:** *White* test for two class comparison and *Kruskal–Wallis* test for multiple class comparisons are used. For both tests the p-value cutoff is set to 0.05;
- **RNA-seq based method:** DESeq function (DESeq2 package) is used by setting “Wald test” as hypothesis testing and the significance cutoff *α* to 0.05. DESeq performs an independent filtering based on the mean of normalized counts for each taxa, optimizing the number of taxa which will have an adjusted p-value (p.adj) below a given false discovery rate (FDR) cutoff (i.e.; alpha). In DESeq, the Benjamini-Hochberg false discovery rate (FDR) is chosen as multiple test correction;
- **Metagenomic based method:** LEfSe (Linear discriminant analysis Effect Size) analysis [68] is carried out at the genus level under the following assumptions: *Wilcoxon* test p-value cutoff is set to 0.05, *Kruskal-Wallis* test p-value cutoff is set to 0.01 and the the Linear Discriminant Analysis (LDA) scores cutoff is set to 4.0. LDA scores represent the effect size of each abundant microbial genera.
- **Supervised learning methods method:** it has been shown that among all the SL methods, the Support Vector Machines (SVMs) can be effectively applied to the microbial community data for classification tasks [69]. Therefore, we have chosen SVMs to identify the features (microbial genera) that my be used to classify samples into PEL confounding factors groups (i.e.; discriminate between groups). The performance of the SVM binary classifier is estimated using the AUC and its graphical representation is provided through the ROC-AUC.

In a case-control study on the microbiota, it is essential to distinguish between disease-related differences and those caused by confounding factors like lifestyle habits or environmental exposures, which can significantly affect microbial composition. Addressing these confounders is necessary to avoid bias in identifying disease markers and prediction modeling [70]. We used a multi-approach strategy to identify microbial taxa associated with potential confounding factors. To ensure the robustness of our findings, we cross-validated results from the Differential Abundance Analysis (DAA). Specifically, taxa associated with Potential Environmental/Lifestyle (PEL) confounding factors, termed “confounded taxa,” were identified if they appeared in the results of at least two DAA methods.

## Results

### Alpha-diversity

We found statistically significant differences in terms of alpha-diversity of microbial community composition only for Nationality and Sport. Pairwise Wilcoxon Rank Sum Tests with Bonferroni correction confirm our findings about alpha-diversity related to Sport and Nationality factors (p-value<=0.05). Figure 1 shows that there are significant differences in terms of richness:

**Figure 1.**
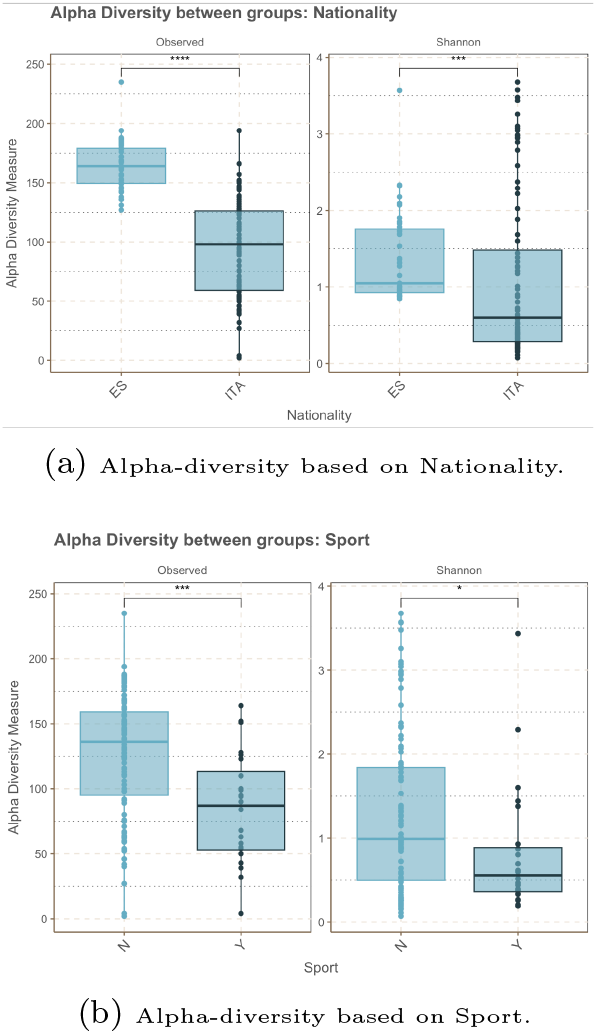
The box plots show the overall differences of microbial community composition based on alpha-diversity. Symbols: ns stands for p-value*>*0.05; * stands for p-value<=0.05; ** stands for p-value<=0.01; *** stands for p-value<=0.001; **** stands for p-value<=0.0001.

- between italian nationality (ITA) subjects and of spanish nationality (ES) subjects (p-value<=0.001 for both Observed and Shannon indexes);
- between subjects who practice sports and those who do not (p-value<=0.001 for observed richness index and p-value<=0.05 for Shannon index).

For Nationality, Figure 1-a) reveals that both indexes, Observed and Shannon assume a higher value for Spanish (ES) then Italian (ITA).

This means that ES subjects group for Nationality is characterized by a more complex microbial profile in term of richness compared to the ITA subjects group. Similarly, N subjects group for Sport is characterized by a higher microbial biodiversity then Y subjects group.

Median values of both Shannon and Observed indexes for Sport and Nationality are reported in the Table 4 and in the Table 5.

**Table 4.**
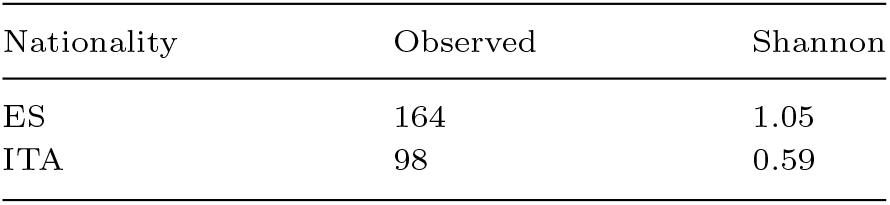
Median values of the Shannon and Observed indexes for Nationality (results are rounded to the second decimal place).

| Nationality | Observed | Shannon |
| --- | --- | --- |
| ES | 164 | 1.05 |
| ITA | 98 | 0.59 |

**Table 5.** Median values of the Shannon and Observed indexes for Sport (results are rounded to the second decimal place).

| Sport | Observed | Shannon |
| --- | --- | --- |
| Y | 136 | 0.99 |
| N | 87 | 0.55 |

**Table 6.** The number of microbial discriminant genera identified by statistical tests is reported for each PEL confounding factor.

| PEL confounding factors | #N microbial discriminant genera |
| --- | --- |
| Sex | 1 |
| Eyeglasses | 7 |
| Computer | 2 |
| Allergy | 4 |
| Sport | 3 |
| Nationality | 2 |
| Bmi | 15 |
| Age | 63 |

### Beta-diversity

Figure 2 shows PCoA analysis results based on the Bray-Curtis dissimilarity. Different shapes or colors in the PCoA diagram represent sample groups. We found statistically significant differences (p-value<=0.05 confirmed by the PERMANOVA test) in terms of beta-diversity of microbial community composition only for Nationality, Age, Sport and Eyeglasses.

**Figure 2.**
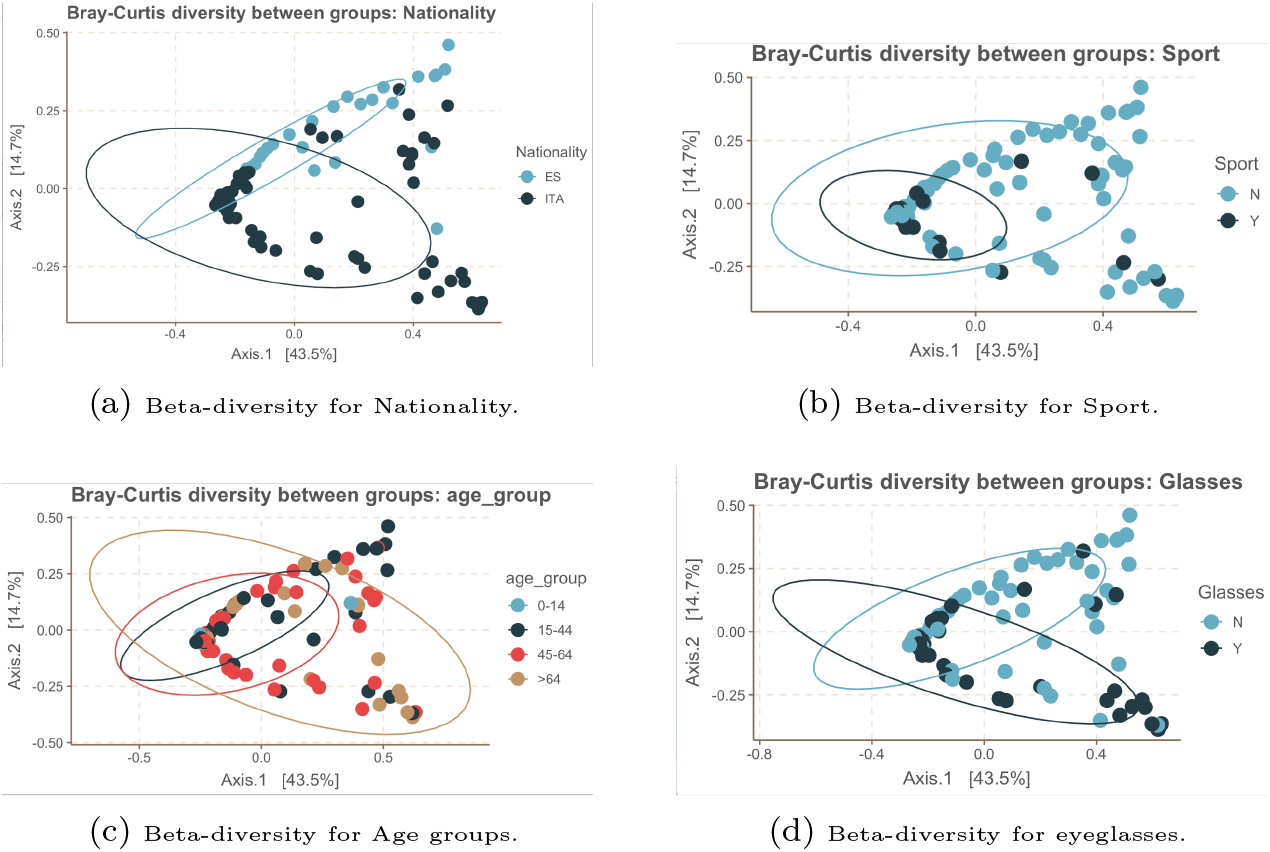
Bray-Curtis distance between samples groups. The ellipses represent the 95% confidence interval of each group.

### Difference Abundance Analysis

#### Statistical (non-parametric) tests

Non-parametric statistical test are applied to identify microbiome taxa at the genus level discriminating between the PEL confounding factors groups. Two-classes comparison is performed by the *White test*, while multi-class comparison is performed by the *Kruskal–Wallis test*. Table 6 shows the number of significant microbial genera (p-value< 0.05) that we found to discriminate between classes for each PEL confounding factor.

#### RNA-seq based method results

Figure 3 and Figure 4 show graphically the DAA results. The *Log2-fold-change* values (y-axis) indicate the differential abundance of the particular genera (x-axis) between the PEL confounding factors classes. DAA results reveal that:

**Figure 3.**
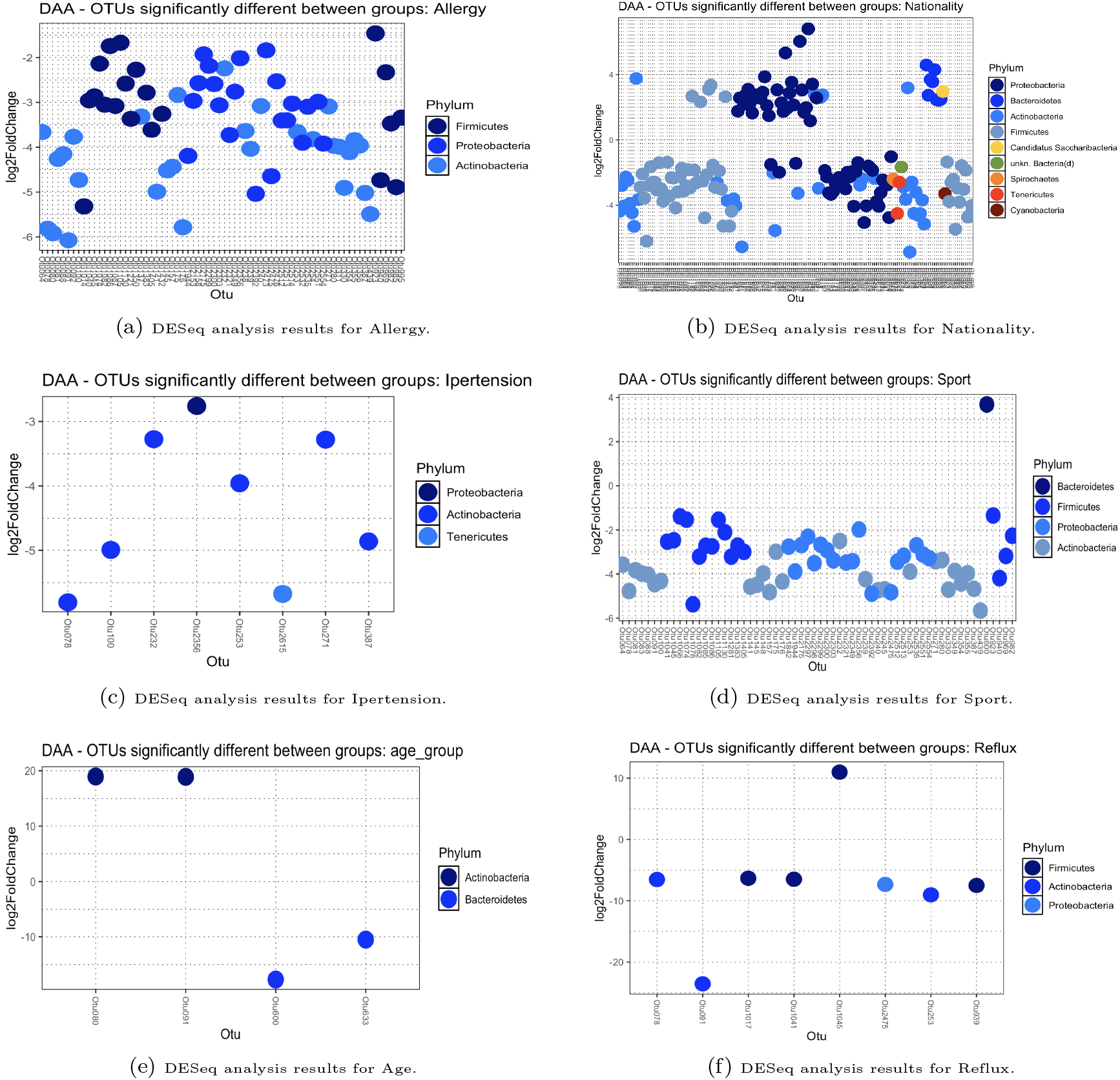
Differences of the OTU abundance between PEL confounding factors groups (Part 1). Results obtained from the DESeq analysis with adjusted p-value cutoff (FDR)<*alpha* are shown (*alpha* is set to 0.01 in DESeq).

**Figure 4.**
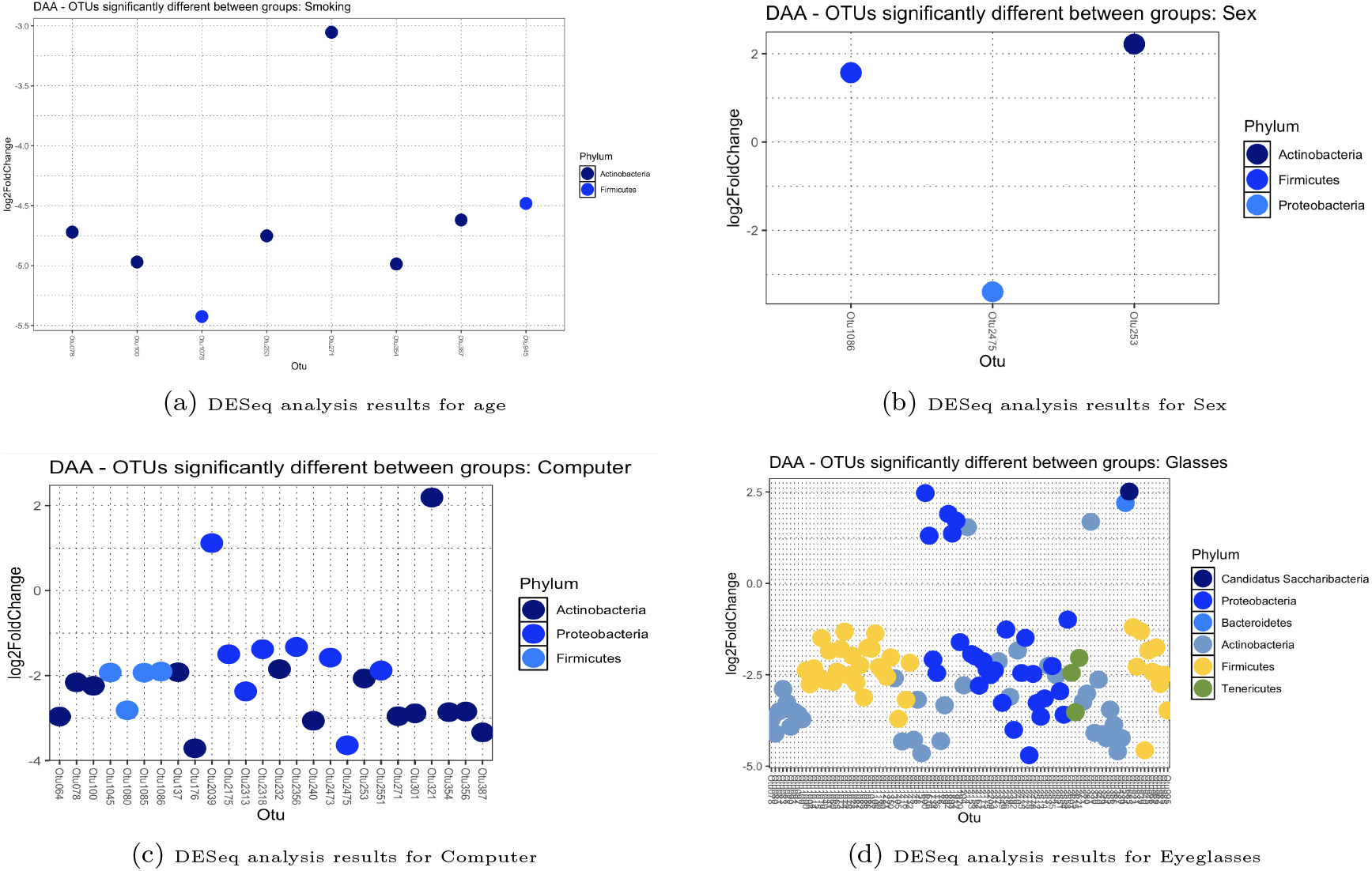
Differences of the OTU abundance between PEL confounding factors groups (Part 2). Results obtained from the DESeq analysis with adjusted p-value cutoff (FDR))<*alpha* are shown (*alpha* is set to 0.01 in DESeq).

- the PEL confounding factors that have the most number (#N> 10) of genera differing in abundance are Allergy, Nationality, Sport, Computer and Eyeglasses;
- the PEL confounding factors that have a number (#N<= 10) of genera differing in abundance are Ipertension, Age, Reflux, Smoking and Sex;
- no difference in abundance is found for BMI.

#### Metagenomic-based method results

Statistically significant differences in the relative abundance of taxa associated with PEL groups are explored using LEfSe. The LEfSe results reveal that there are several microbial genera that differ in abundances for six PEL confounding factors. Table 7 shows the number of discriminant genera identified for each PEL confounding factor. For the rest of the PEL confounding factors no discriminant genera is identified as significant. Figure 5 shows the same LEfSe results in a graphical way.

**Table 7.** The number of microbial discriminant genera identified by LEfSe for each PEL confounding factor is reported. Only the results statistically significant (LDA score> 4.0) are reported in the table.

| PEL confounding factors | #N microbial discriminant genera |
| --- | --- |
| eyeglasses | 7 |
| computer | 4 |
| allergy | 3 |
| sport | 4 |
| nationality | 13 |
| ipertension | 13 |

**Figure 5.**
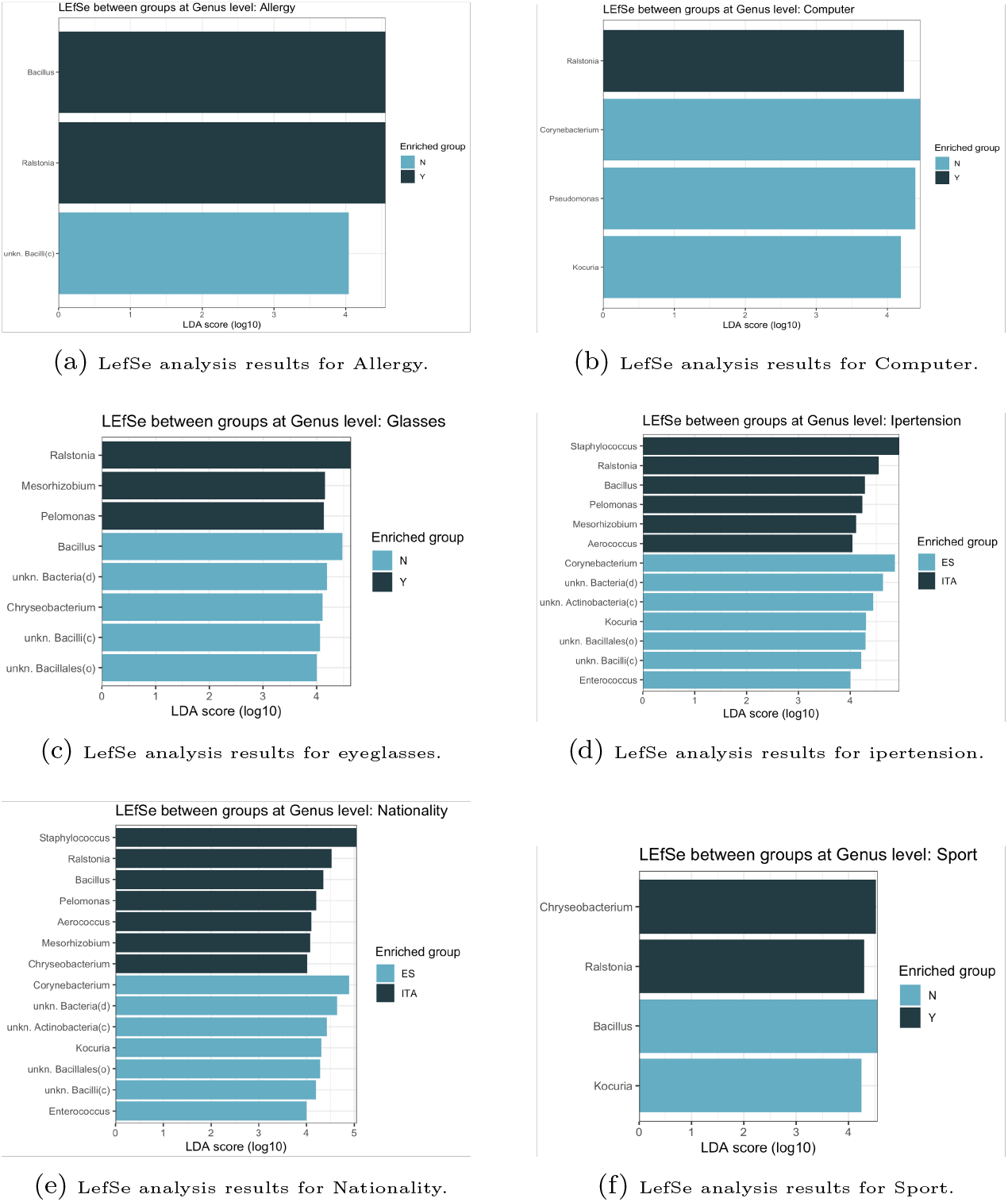
These figures show the genera that differ in abundance between PEL confounding classes. The LDA score is shown at logarithmic scale underneath the bars (LDA score> 4.0).

#### Supervised learning method results

We used the Table 8 as a reference for the quality rating of the acceptable SVM models. In this study, we consider acceptable the classifiers with AUC-ROC score≥0.8 [71] and a confidence interval of AUC equal to 95% [72, 73, 74]. Figure 6 shows the ROC curve and the AUC value for the Nationality. For the other PEL confounding factors AUC-ROC score is <0.8 and therefore it is not shown. Each point on the ROC curve represents the true positive rate (TPR) vs the false positive rate (FPR). The TPR (a.k.a. sensitivity) is the proportion of observations that are correctly predicted to be positive out of all positive observations. Similarly, the FPR (a.k.a. specificity) is the proportion of observations that are incorrectly predicted to be positive out of all negative observations:

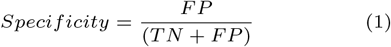

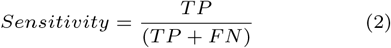

**Table 8.** Quality rating of the acceptable classifiers(i.e.; AUC-ROC≥0.8).

| AUC-ROC score | Quality |
| --- | --- |
| $0.9 \geq \text{score}$ | Excellent |
| $0.8 \geq \text{score} < 0.9$ | Good |

**Figure 6.**
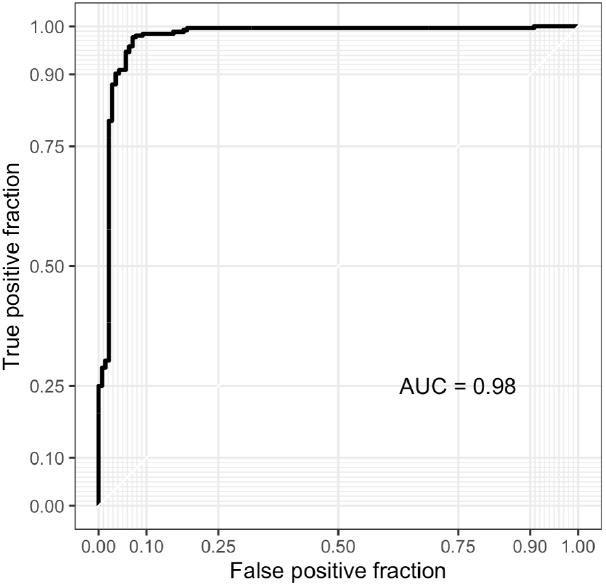
ROC curve and AUC value for Nationality.

## Cross-Validation of the Difference

### Abundance Analysis Results

Confounded taxa can be used to discriminate the microbial communities between classes of a single o multiple communities covary with the PEL confounding factors. Our analysis revealed noteworthy findings regarding the identification of confounded taxa associated with Potential Environmental/Lifestyle (PEL) factors. Specifically, the Venn diagrams presented in Figure 7 show the commonalities and differences between the results sets achieved as output from the different four methods that we applied.

**Figure 7.**
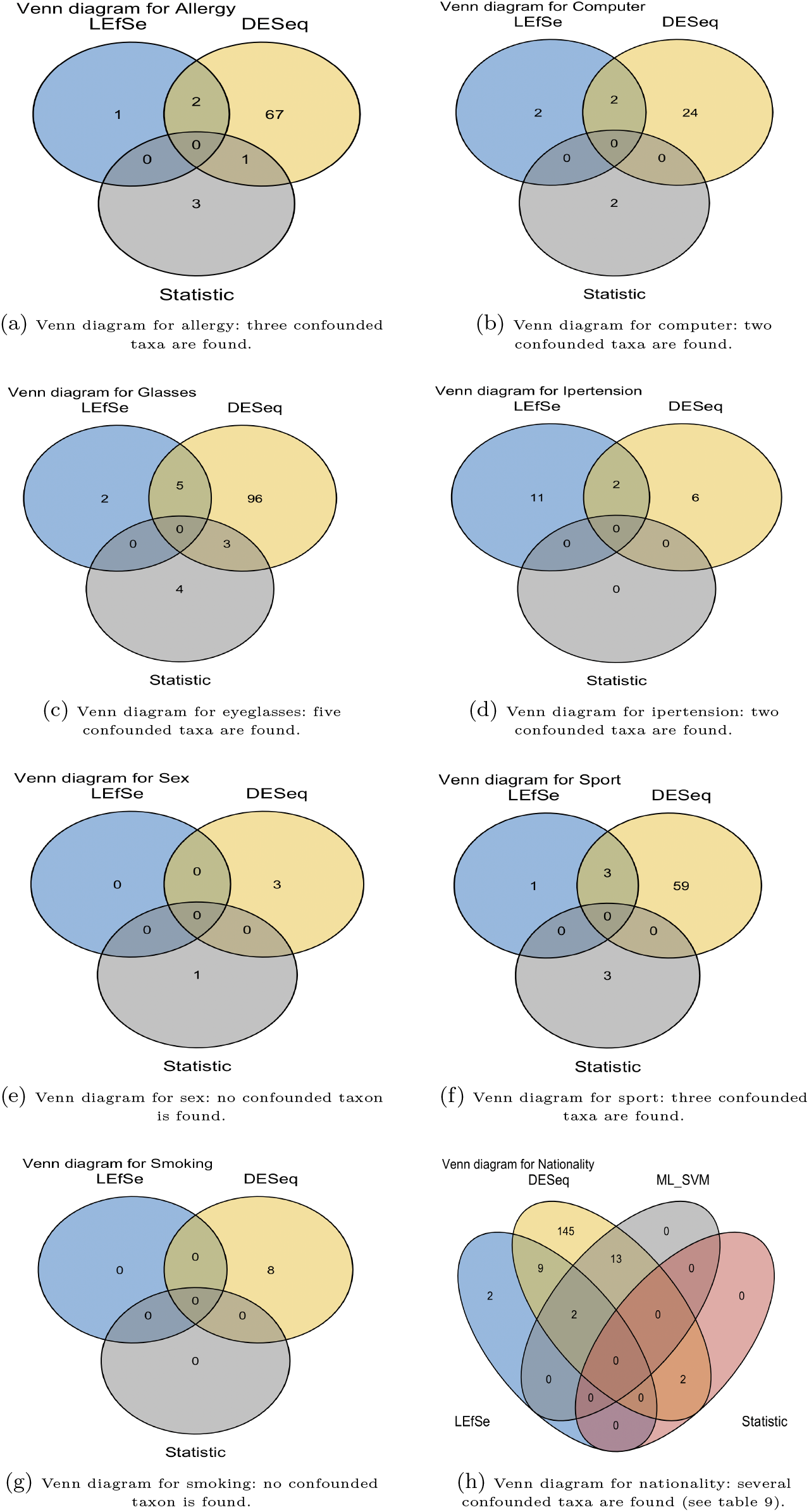
Venn diagrams show the confounded taxa discovered for each PEL confounding factor. Each circle represent the results set of a DAA method (i.e.; “Statistic”,”LEfSe”, “DESeq”, “SVM”).

Remarkably, our SVM-based classifier achieved acceptable performance (AUC-ROC score ≥ 0.8) solely for the Nationality factor. Consequently, the Venn diagram in Figure 7-h) shows four circles for Nationality, unlike the others which show only three.

In particular, our analysis indicates that Nationality exhibited the highest number of confounded taxa as shown in the table 9. In particular, for Nationality 15 genera are found by SVM and DESeq methods, 2 genera are found by SVM, DESeq and LEfSeq methods, 2 genera are found by DESeq and statistic methods and 11 genera are found by DESeq and LEfSeq methods. These confounded taxa hold significance as they can potentially partition the population based on geographic origins.

**Table 9.**
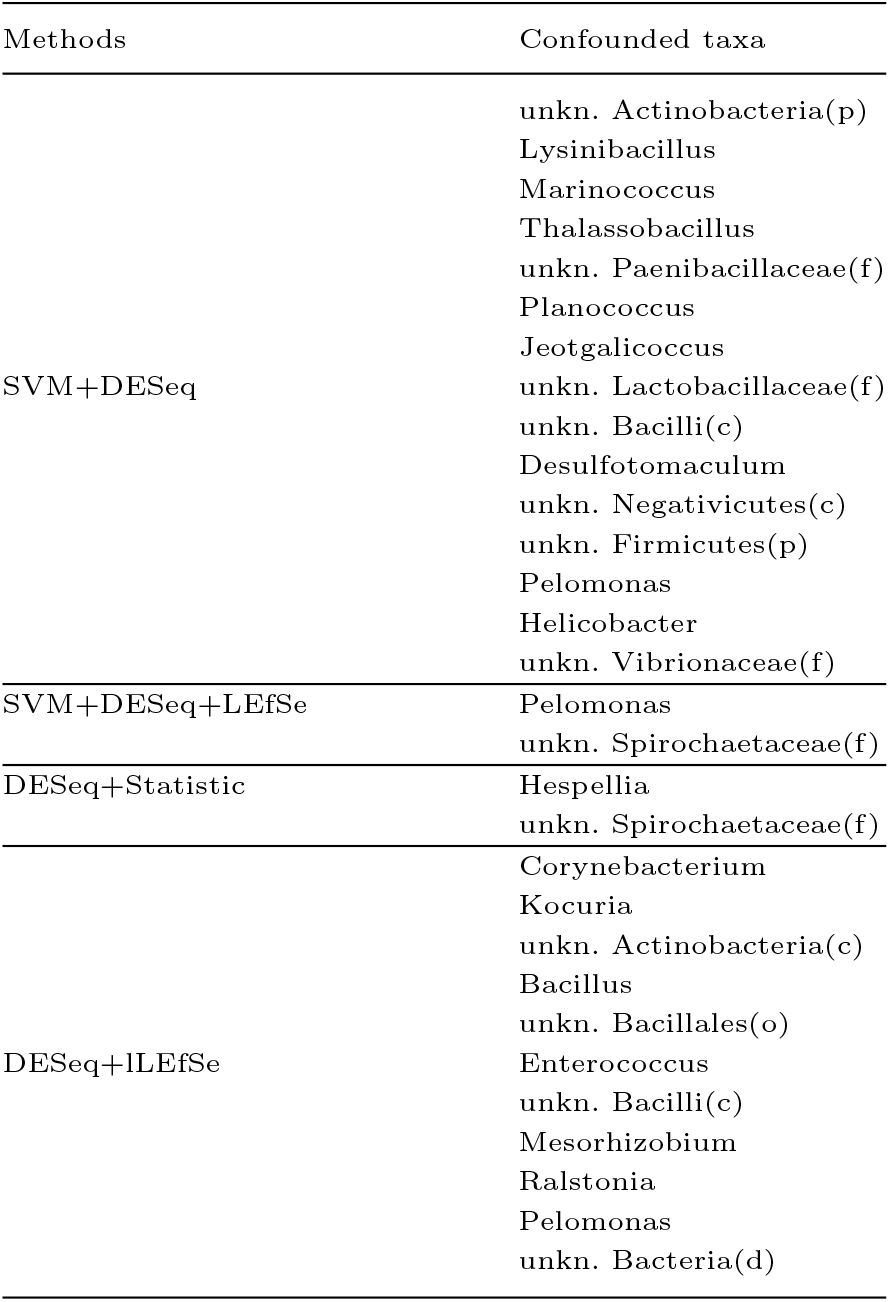
Table shows the confounded taxa discovered for the Nationality. The first column indicates the methods that are applied, the second column lists the confounded taxa found by the corresponding methods in the first column.

## Discussion

As highlighted in the introduction, there has been a noticeable surge in interest in microbiome analysis in recent years. This interest spans various medical fields, encompassing studies on both healthy and unhealthy individuals. Initially, the focus was primarily on understanding how microbiome diversity affects the progression to diseases, particularly exploring outcomes and influencing factors that were previously challenging to discern using available analytical methods [75].

Traditionally, the analysis mainly targeted the microbiome diversity in human feces due to its practicality and direct relationship with microbial populations. Numerous studies have evaluated its significance in estimating disease severity, progression, and the intricate interactions between humans and various biological agents.

However, more recent advancements have revealed the presence of microbiomes in previously considered sterile areas of the body, like the pulmonary microbiome, opening new avenues for research and potential therapeutic interventions [76]. Moreover, emerging evidence has shed light on the microbiome’s specific role in influencing widespread diseases such as endometriosis, particularly in its interaction with other pathogenic bacteria, and further data may clarify this relationship.

Furthermore, it’s crucial to acknowledge that different microbiome populations communicate across bodily systems, despite their spatial separation, potentially exerting cross-system influences. Notably, there’s growing interest in how the gut microbiome might impact ocular surface homeostasis and associated disorders [77, 78, 79].

Our study represents one of the initial efforts to delve deeper into microbial diversity within the eyes of healthy individuals, utilizing innovative techniques [80]. This research carries significant implications as its findings can inform future investigations in both healthy and unhealthy populations. Importantly, our methodology incorporates measures to mitigate factors that could distort results, such as meticulous sampling techniques and patient selection, thereby enhancing its external validity, and applied methodology, with a multi-level approach. On top of this, data processing has been reviewed thoroughly in order to limit the uninformative OTUs, as described in the previous sections.

First step methodologies were applied in order to calculate Alpha and Beta-diversity. Advanced methods were applied in order to describe the confounding factors. Our research highlights how environmental and lifestyle factors, such as Nationality and Sport, significantly influence the ocular surface microbiome’s composition. These findings could have important implications for understanding microbial diversity and its relationship with environmental and behavioral variables.

### Alpha-diversity

Analyzing alpha diversity, we found statistically significant differences for Nationality and Sport. Specifically, we observed: A higher microbial richness in Spanish subjects compared to Italian subjects, as described by Observed and Shannon indexes. A higher microbial biodiversity in subjects who practice sports compared to those who do not for the observed richness index and for the Shannon index. These results suggest that the group of Spanish subjects has a more complex microbial profile in terms of richness compared to Italian subjects and that sports practitioners exhibit greater microbial biodiversity than non-practitioners (Figure 1). Previous studies have shown that geographic location can significantly impact the composition of the ocular surface microbiome [81]. For example, the ocular surface metagenome of young Han Chinese from Beijing, Wenzhou, and Guangzhou, was examined demonstrating distinct microbial communities shaped by geographic differences [29]. This highlights the importance of considering geographic variation when studying the ocular microbiome.

Additionally, other studies provided a molecular characterization of the ocular microbiome, noting that it is a unique and low microbial environment influenced by various external factors, including geographic location [80]. This molecular insight supports our findings that nationality, as a proxy for geographic and environmental differences, significantly impacts the microbial diversity and composition of the ocular surface.These data should be confirmed in future studies assessing the same variables with respect to other nationalities. Additionally, it should be noted that in this analysis the Alpha diversity from the OS was generally higher than previous reports [4]. Conversely, contradictory findings exist regarding the relationship between microbial diversity and physical activity. While some studies suggest a positive correlation, others propose that increased microbial diversity may not always signify a healthier state due to potential proliferation of harmful bacteria.

### Beta-Diversity

On the other hand, when it comes to beta diversity, we found statistically significant differences in microbial community composition concerning Nationality, Age, Sport, and Eyeglasses use. Moreover, when analyzing differential abundance, Non-parametric statistical tests were applied to identify microbiome taxa at the genus level discriminating between PEL confounding factors groups. The results show: The highest number of differential microbial genera for Allergy, Nationality, Sport, Computer use, and Eyeglasses use. A lower number of differential genera for Hypertension, Age, Reflux, Smoking, and Sex. No significant difference for BMI. This is particularly relevant in view of the fact that beta-diversity describes the inter-individual variability, and may describe trajectories over time and space (Figure 2). This result also gives value to the taxonomic profiles from the samples, and confirms data from previous studies in different group populations [36].

For future research, these variables should be considered attentively when evaluating biomarkers or microbiome composition, as they showed to “cluster” around PEL that may describe patterns in study populations, e.g. lower number of differential genera for variables that are more commonly encountered in older persons age, hypertension- and higher number of differential general in possibly younger persons - sport, computer, although that is not widely ascertained.

Curiously, the finding that no significant differences were found with respect to BMI do not align with observations from other studies [82]. No comparison is possible with other data from eyeglass use, as no data have been retrieved from our literature search. For what pertains to gender, previous studies have suggested that this may also play a significant role in the composition of the ocular surface microbiome. For instance, some authors discussed the broader impact of the microbiome on ocular surface pathophysiology and disorders, emphasizing potential gender differences in microbial composition [83, 84, 81]. These findings underscore the influence of demographic and lifestyle factors on the ocular microbiome. Notably, the PERMANOVA test confirmed the statistical significance of these differences, highlighting the robustness of our result.

### Difference Abundance Analysis

When it comes to the DAAs, microbial discriminant genera were identified for each PEL factor, showing high variability (from 1 discriminant for Sex to 63 for Age see Table 6). PEL factors with above or below 10 genera differing in abundance show relevant differences. Furthermore, this analysis is confirmed by metagenomic based method results (Figure 5). In particular, Using LEfSe, we explored statistically significant differences in the relative abundance of taxa associated with PEL groups. The results revealed significant differences for six PEL confounding factors, with varying numbers of discriminant genera identified for each factor. Machine learning-based approaches provided valuable insights into feature selection and classification tasks. However, the relatively small sample size in our cohort (135 samples) compared to the high dimensionality of the dataset (2668 OTUs) necessitates cautious interpretation of the results. As already outlined, our SVM-based classifier achieved acceptable performance solely for the Nationality factor. While supervised learning methods offer promising avenues for predictive modeling, their outcomes should be corroborated by other analytical techniques to ensure robustness and generalizability. More importantly, Venn diagram and other methods were concordant in identifying the considered variables. This highlights how our methodology is for concordance between methods (7, 9), supporting the validity of our approach across methods. In this sense, it would be interesting to verify these data including more nationality in a multicenter study. Ultimately, in this study we could not directly assess the eye community state type (ECST) that was previously described [4], as the aim of the analysis was different and a different descriptive level would have been required, including additional analysis.

### Cross-Validation of the Difference Abundance Analysis Results

Our cross-validation approach aimed to mitigate the impact of confounding factors on microbial composition analysis. By identifying taxa consistently associated with potential environmental/lifestyle (PEL) confounding factors, we elucidated their role in shaping microbial communities. Notably, taxa correlated with nationality, sport, and eyeglasses usage provide valuable insights into the geographical and lifestyle determinants of ocular microbiome diversity. Geographical factors represent a complex interplay of genetic, environmental, and cultural influences on microbial composition.

Consistent with findings in gut microbiome research, our study revealed geographical variations in the ocular microbiome, emphasizing the need for further investigation into the underlying mechanisms driving these differences [85, 60, 59, 86]. Moreover, our analysis identified sport and eyeglasses usage as potential determinants of ocular microbiome composition. Emerging evidence suggests a correlation between physical activity and gut microbiota composition, raising intriguing possibilities regarding its impact on the ocular microbiome [70, 41, 40].

Similarly, while contact lenses have been implicated in ocular infections, the association between eyeglasses and ocular microbiome remains relatively unexplored [87, 88, 89]. In summary, our findings shed light on the complex interplay between environmental, lifestyle, and demographic factors in shaping the ocular microbiome of healthy individuals. Further research is warranted to unravel the underlying mechanisms and implications for ocular health management and personalized medicine

## Conclusion

Our study underscores the significant impact of various environmental and lifestyle factors on the ocular surface microbiome. Factors such as nationality, sport, age, smoking, and computer use play critical roles in shaping microbial diversity and composition. Future research should further explore these interactions to develop personalized approaches for managing ocular health. It is essential to consider these factors to achieve unbiased conclusions about the microbiome’s association with human diseases. By understanding these influences, we can better identify differences related to diseases and improve strategies for ocular health management and disease prevention. The findings of this study have to be seen in light of a potential limitation due to the size of the participant cohort. Small sample size can indeed introduce a limited generalizability of the study findings. This limitation could be overcome in a future work by a multi-center research study that involves several independent medical institutions in enrolling more participants to increase the sample size.

## Data Availability

All data produced in the present study are available upon reasonable request to the authors

## Data and availability

Data are available from the corresponding authors upon reasonable request.

## Author contributions

M.S. and V.R. contributed to the study conception and design; M.S. performed the bioinformatics analysis and wrote the manuscript; G.C., G.S., D.B., M.N., J.V., R.M., G.L. and V.R. contributed to the interpretation of the results, reviewed and wrote the manuscript; C.A., V.R., D.B., C.R.L., C.M. and M.R. provided the dataset; F.R. and G.G. supervised the study design. All authors reviewed the results and approved the final version of the manuscript.

## Acknowledgments

This work was supported by the Italian Ministry of Health “Fondi Ricerca Corrente” to IRCCS Sacro Cuore Don Calabria Hospital.

We thank Sequentia Biotech SL (Barcelona, Spain) for the support to the project and the analysis of the data.

## References

1. Jane Peterson, Susan Garges, Maria Giovanni, Pamela McInnes, Lu Wang, Jeffery A Schloss, Vivien Bonazzi, Jean E McEwen, Kris A Wetterstrand, Carolyn Deal, et al. The nih human microbiome project. Genome research, 19(12):2317–2323, 2009.

2. Structure, function and diversity of the healthy human microbiome. nature, 486(7402):207–214, 2012.

3. Kara M Cavuoto, Santanu Banerjee, and Anat Galor. Relationship between the microbiome and ocular health. The ocular surface, 17(3):384–392, 2019.

4. Davide Borroni, Andreu Paytuví Gallart, Walter Sanseverino, Carmen Gómez-Huertas, Paola Bonci, Vito Romano, Giuseppe Giannaccare, Miguel Rechichi, Alessandro Meduri, Giovanni Oliverio, and Carlos Rocha de Lossada. Exploring the healthy eye microbiota niche in a multicenter study. International Journal of Molecular Sciences, 23:10229, 09 2022.

5. O. Paliy and B. D. Foy. Mathematical modeling of 16S ribosomal DNA amplification reveals optimal conditions for the interrogation of complex microbial communities with phylogenetic microarrays. Bioinformatics, 27(15):2134–2140, Aug 2011.

6. Zhe Luan, Shihui Fu, Shirui Qi, Congyong Li, Jun Chen, Yiming Zhao, Hanwen Zhang, Junling Wu, Zhizhuang Zhao, Jiaqi Zhang, Yi Chen, Wei Zhang, Yujia Jing, Shufang Wang, and Gang Sun. A metagenomics study reveals the gut microbiome as a sex-specific modulator of healthy aging in hainan centenarians. Experimental Gerontology, 186:112356, 2024.

7. A. K. Whitehead, M. C. Meyers, C. M. Taylor, M. Luo, S. E. Dowd, X. Yue, and L. O. Byerley. Sex-Dependent Effects of Inhaled Nicotine on the Gut Microbiome. Nicotine Tob Res, 24(9):1363–1370, Aug 2022.

8. W. M. de Vos, H. Tilg, M. Van Hul, and P. D. Cani. Gut microbiome and health: mechanistic insights. Gut, 71(5):1020–1032, May 2022.

9. A. Matysiak, M. Kabza, J. A. Karolak, M. M. Jaworska, M. Rydzanicz, R. Ploski, J. P. Szaflik, and Gajecka. Characterization of Ocular Surface Microbial Profiles Revealed Discrepancies between Conjunctival and Corneal Microbiota. Pathogens, 10(4), Mar 2021.

10. Davide Borroni, Vito Romano, Stephen Kaye, Tobi Somerville, Luca Napoli, Adriano Fasolo, Paola Gallon, Diego Ponzin, Alfonso Esposito, and Stefano Ferrari. Metagenomics in ophthalmology: Current findings and future prospectives. BMJ open ophthalmology, 4, 06 2019.

11. Heleen Delbeke, Saif Younas, Ingele Casteels, and Marie Joossens. Current knowledge on the human eye microbiome: A systematic review of available amplicon and metagenomic sequencing data. Acta Ophthalmologica, 2020.

12. Yutong Kang, Shudan Lin, Xueli Ma, Yanlin Che, Yi–Ju Chen, Tian Wan, Die Zhang, Jiao Shao, Jie Xu, Yi Xu, Yongliang Lou, and Meiqin Zheng. Strain heterogeneity, cooccurrence network, taxonomic composition and functional profile of the healthy ocular surface microbiome. Eye and Vision, 2021.

13. Sisinthy Shivaji, Rajagopalaboopathi Jayasudha, Gumpili Sai Prashanthi, Sama Kalyana Chakravarthy, and Savitri Sharma. The human ocular surface fungal microbiome, 2019.

14. Thanachaporn Kittipibul and Vilavun Puangsricharern. The ocular microbiome in stevens-johnson syndrome, 2021.

15. Davide Borroni, Carlos Rocha de Lossada, Cosimo Mazzotta, Jose-María Sánchez-González, Filomena Papa, and Federico Gabrielli. Ocular microbiome evaluation in dry eye disease and meibomian gland dysfunction: Values of variables. Experimental Eye Research, pages 109656–109656, 2023.

16. Carlos Rocha-de Lossada, Cosimo Mazzotta, Federico Gabrielli, Filomena Tiziana Papa, Carmen Gómez-Huertas, Celia García-López, Facundo Urbinati, Rahul Rachwani-Anil, María García-Lorente, José-María Sánchez-González, et al. Ocular surface microbiota in naïve keratoconus: A multicenter validation study. Journal of Clinical Medicine, 12(19):6354, 2023.

17. Domenico Schiano-Lomoriello, Irene Albicca, Laura Contento, Federico Gabrielli, Cinzia Alfonsi, Fabio Pietro, Filomena Papa, Antonio Ballesteros-Sánchez, José-María Sánchez-González, Carlos Rocha de Lossada, Cosimo Mazzotta, Giuseppe Giannaccare, Chiara Bonzano, and Davide Borroni. Infectious keratitis: Characterization of microbial diversity through species richness and shannon diversity index. Biomolecules, 14:389, 03 2024.

18. Carlos Rocha de Lossada, Cosimo Mazzotta, Federico Gabrielli, Filomena Papa, Carmen Gómez-Huertas, Celia García-López, Facundo Urbinati, Rahul Rachwani-Anil, Maria Garcia Lorente, José-María Sánchez-González, Miguel Rechichi, Giovanni Rubegni, and Davide Borroni. Ocular surface microbiota in naïve keratoconus: A multicenter validation study. Journal of Clinical Medicine, 12:6354, 10 2023.

19. Yusen Huang, Bingbing Yang, and W. Li. Defining the normal core microbiome of conjunctival microbial communities, 2016.

20. Jerome Ozkan, Shaun Nielsen, Cristina Diez-Vives, Minas Coroneo, Torsten Thomas, and Mark Willcox. Temporal stability and composition of the ocular surface microbiome. Scientific reports, 7(1):9880, 2017.

21. Qunfeng Dong, Jennifer M Brulc, Alfonso Iovieno, Brandon Bates, Aaron Garoutte, Darlene Miller, Kashi V Revanna, Xiang Gao, Dionysios A Antonopoulos, Vladlen Z Slepak, et al. Diversity of bacteria at healthy human conjunctiva. Investigative ophthalmology & visual science, 52(8):5408–5413, 2011.

22. Yutong Kang, Hao Zhang, Meina Hu, Yao Ma, Pengfei Chen, Zelin Zhao, Jinyang Li, Yuee Ye, Meiqin Zheng, and Yongliang Lou. Alterations in the ocular surface microbiome in traumatic corneal ulcer patients. Investigative Opthalmology & Visual Science, 2020.

23. Pachiappan Arjunan and Radhika Swaminathan. Do oral pathogens inhabit the eye and play a role in ocular diseases?, 2022.

24. Kara M. Cavuoto and Santanu Banerjee. Anatomic characterization of the ocular surface microbiome in children, 2019.

25. Fiona Stapleton, Ajay Kumar Vijay, and Nicole Carnt. Epidemiology, microbiology, and genetics of contact lens–related and non–contact lens-related infectious keratitis, 2022.

26. Jennifer Wing-Ki Yau, Jianbo Hou, Stephen Kwok-Wing Tsui, Ting Fan Leung, Nam Sze Cheng, Jason C. Yam, Ka Wai Kam, Vishal Jhanji, and Kam Lun Hon. Characterization of ocular and nasopharyngeal microbiome in allergic rhinoconjunctivitis, 2019.

27. Michael E. Zegans and Russell N. Van Gelder. Considerations in understanding the ocular surface microbiome, 2014.

28. Kent A. Willis, Cameron Postnikoff, Amelia Freeman, Gabriel Rezonzew, Kelly K. Nichols, Amit Gaggar, and Charitharth Vivek Lal. The closed eye harbors a unique microbiome in dry eye disease, 2020.

29. Yuhua Deng, Xiaofeng Wen, Xiao Hu, Yanli Zou, Chan Zhao, Xue-Jiao Chen, Miao Li, Xifang Li, Xiaoyi Deng, Paul W. Bible, Hongmin Ke, Jiahao Situ, Shixin Guo, Juanran Liang, Tingting Chen, Bin Zou, Yu Liu, Wei Chen, Kaili Wu, Meifen Zhang, Zi-Bing Jin, Lingyi Liang, and Lai Wei. Geographic difference shaped human ocular surface metagenome of young han chinese from beijing, wenzhou, and guangzhou cities, 2020.

30. Y. Wang, H. Chen, Tian Xia, and Yusen Huang. Characterization of fungal microbiota on normal ocular surface of humans, 2020.

31. Yutong Kang, Shudan Lin, Xueli Ma, Yanlin Che, Yi–Ju Chen, Tian Wan, Die Zhang, Jiao Shao, Jie Xu, Yi Xu, Yongliang Lou, and Meiqin Zheng. Strain heterogeneity, cooccurrence network, taxonomic composition and functional profile of the healthy ocular surface microbiome, 2021.

32. Davide Borroni, Andreu Paytuví-Gallart, Walter Sanseverino, Carmen Gómez-Huertas, Paola Bonci, Vito Romano, Giuseppe Giannaccare, Miguel Rechichi, Alessandro Meduri, Giovanni William Oliverio, et al. Exploring the healthy eye microbiota niche in a multicenter study. International Journal of Molecular Sciences, 23(18):10229, 2022.

33. Glenn W. Suter II and Susan M. Cormier. A method for assessing the potential for confounding applied to ionic strength in central appalachian streams. Environmental Toxicology and Chemistry, 32(2):288–295, 2013.

34. Pasquale Aragona, Christophe Baudouin, Jose Castillo, Elisabeth Messmer, Stefano Barabino, Jesús Merayo-Lloves, Francoise Brignole-Baudouin, Leandro Inferrera, Maurizio Rolando, Rita Mencucci, Maria Rescigno, Stefano Bonini, and Marc Labetoulle. The ocular microbiome and microbiota and their effects on ocular surface pathophysiology and disorders. Survey of Ophthalmology, 66, 04 2021.

35. Yanjiao Zhou, Martin Holland, Pateh Makalo, Hassan Joof, Chrissy Roberts, David Mabey, Robin Bailey, Matthew Burton, George Weinstock, and Sarah Burr. The conjunctival microbiome in health and trachomatous disease: A case control study. Genome medicine, 6:99, 11 2014.

36. X. Wen, L. Miao, Y. Deng, P. W. Bible, X. Hu, Y. Zou, Y. Liu, S. Guo, J. Liang, T. Chen, G. H. Peng, W. Chen, L. Liang, and L. Wei. The Influence of Age and Sex on Ocular Surface Microbiota in Healthy Adults. Invest Ophthalmol Vis Sci, 58(14):6030–6037, Dec 2017.

37. Melissa Aldridge and R Sean Morrison. Study design, precision, and validity in observational studies. Journal of palliative medicine, 12:77–82, 02 2009.

38. Braden T. Tierney, Yingxuan Tan, Zhen Yang, Bing Shui, Michaela J. Walker, Benjamin M. Kent, Aleksandar D. Kostic, and Chirag J. Patel. Systematically assessing microbiome–disease associations identifies drivers of inconsistency in metagenomic research. PLOS Biology, 20(3):1–18, 03 2022.

39. E. Gagnon, P. L. Mitchell, H. D. Manikpurage, cE. Abner, N. Taba, T. Esko, N. Ghodsian, S. riault, P. Mathieu, and B. J. Arsenault. Impact of the gut microbiota and associated metabolites on cardiometabolic traits, chronic diseases and human longevity: a Mendelian randomization study. J Transl Med, 21(1):60, Jan 2023.

40. A. Ahmad, M. Imran, and H. Ahsan. Biomarkers as Biomedical Bioindicators: Approaches and Techniques for the Detection, Analysis, and Validation of Novel Biomarkers of Diseases. Pharmaceutics, 15(6), May 2023.

41. Darya Chyzhyk, Gaël Varoquaux, Michael Peter Milham, and Bertrand Thirion. How to remove or control confounds in predictive models, with applications to brain biomarkers. GigaScience, 11, 2022.

42. S. Weiss, Z. Z. Xu, S. Peddada, A. Amir, K. Bittinger, A. Gonzalez, C. Lozupone, J. R. Zaneveld, Y. zquez Baeza, A. Birmingham, E. R. Hyde, and R. Knight. Normalization and microbial differential abundance strategies depend upon data characteristics. Microbiome, 5(1):27, Mar 2017.

43. S. Wang. Multiscale adaptive differential abundance analysis in microbial compositional data. Bioinformatics, 39(4), Apr 2023.

44. Marzia Settino and Mario Cannataro. Mmrfbiolinks: an r-package for integrating and analyzing mmrf-commpass data. Briefings in bioinformatics, 22, 04 2021.

45. Marzia Settino and Mario Cannataro. Mmrfvariant: Prioritizing variants in multiple myeloma. Informatics in Medicine Unlocked, 39:101271, 2023.

46. Tiantian Liu, Chao Zhou, Huimin Wang, Hongyu Zhao, and Tao Wang. phylomda: an r package for phylogeny-aware microbiome data analysis. BMC Bioinformatics, 23, 06 2022.

47. N. Segata, J. Izard, L. Waldron, D. Gevers, L. Miropolsky, W. S. Garrett, and C. Huttenhower. Metagenomic biomarker discovery and explanation. Genome Biol, 12(6):R60, Jun 2011.

48. Michelle Hagen, Rupashree Dass, Cathy Westhues, Jochen Blom, Sebastian Schultheiss, and Sascha Patz. Interpretable machine learning decodes soil microbiome’s response to drought stress. 12 2023.

49. M. Calgaro, C. Romualdi, L. Waldron, D. Risso, and N. Vitulo. Assessment of statistical methods from single cell, bulk RNA-seq, and metagenomics applied to microbiome data. Genome Biol, 21(1):191, Aug 2020.

50. D. Knights, E. K. Costello, and R. Knight. Supervised classification of human microbiota. FEMS Microbiol Rev, 35(2):343–359, Mar 2011.

51. D. Knights, J. Kuczynski, O. Koren, R. E. Ley, D. Field, R. Knight, T. Z. DeSantis, and S. T. Kelley. Supervised classification of microbiota mitigates mislabeling errors. ISME J, 5(4):570–573, Apr 2011.

52. J. T. Nearing, G. M. Douglas, M. G. Hayes, J. MacDonald, D. K. Desai, N. Allward, C. M. A. Jones, R. J. Wright, A. S. Dhanani, A. M. Comeau, and M. G. I. Langille. Author Correction: Microbiome differential abundance methods produce different results across 38 datasets. Nat Commun, 13(1):777, Feb 2022.

53. Paul J. McMurdie and Susan Holmes. phyloseq: An r package for reproducible interactive analysis and graphics of microbiome census data. PLOS ONE, 8(4):1–11, 04 2013.

54. Yang Cao, Qingyang Dong, Dan Wang, Pengcheng Zhang, Ying Liu, and Chao Niu. microbiomeMarker: an R/Bioconductor package for microbiome marker identification and visualization. Bioinformatics, 38(16):4027–4029, 06 2022.

55. M. I. Love, W. Huber, and S. Anders. Moderated estimation of fold change and dispersion for RNA-seq data with DESeq2. Genome Biol, 15(12):550, 2014.

56. Yutong Kang, Leihao Tian, Xiaobin Gu, Yiju Chen, Xueli Ma, Shudan Lin, Zhenjun Li, Yongliang Lou, and Meiqin Zheng. Characterization of the ocular surface microbiome in keratitis patients after repeated ophthalmic antibiotic exposure. Microbiology Spectrum, 10(2):e02162–21, 2022.

57. Hakdong Shin, Kenneth Price, Luong Albert, Jack Dodick, Lisa Park, and Maria Gloria Dominguez-Bello. Changes in the eye microbiota associated with contact lens wearing. MBio, 7(2):10–1128, 2016.

58. Yanlin Zhong, Xie Fang, Xuemei Wang, Yu-An Lin, Huping Wu, and Cheng Li. Effects of sodium hyaluronate eye drops with or without preservatives on ocular surface bacterial microbiota. Frontiers in Medicine, 9:793565, 2022.

59. Wei Dai, Cai Li, Ting Li, Jianchang Hu, and Heping Zhang. Super-taxon in human microbiome are identified to be associated with colorectal cancer. BMC Bioinformatics, 23, 06 2022.

60. S. ndez, T. ý, and P. Baldrian. The concept of operational taxonomic units revisited: genomes of bacteria that are regarded as closely related are often highly dissimilar. Folia Microbiol (Praha), 64(1):19–23, Jan 2019.

61. S. Weiss, Z. Z. Xu, S. Peddada, A. Amir, K. Bittinger, A. Gonzalez, C. Lozupone, J. R. Zaneveld, Y. zquez Baeza, A. Birmingham, E. R. Hyde, and R. Knight. Normalization and microbial differential abundance strategies depend upon data characteristics. Microbiome, 5(1):27, Mar 2017.

62. M. Nikodemova, E. A. Holzhausen, C. L. Deblois, J. H. Barnet, P. E. Peppard, G. Suen, and K. M. Malecki. The effect of low-abundance OTU filtering methods on the reliability and variability of microbial composition assessed by 16S rRNA amplicon sequencing. Front Cell Infect Microbiol, 13:1165295, 2023.

63. Y. Hu, G. A. Satten, and Y. J. Hu. LOCOM: A logistic regression model for testing differential abundance in compositional microbiome data with false discovery rate control. Proc Natl Acad Sci U S A, 119(30):e2122788119, Jul 2022.

64. M. D. Robinson and A. Oshlack. A scaling normalization method for differential expression analysis of RNA-seq data. Genome Biol, 11(3):R25, 2010.

65. Tao Wen, Guoqing Niu, Tong Chen, Qirong Shen, Jun Yuan, and Yong-Xin Liu. The best practice for microbiome analysis using R.

66. F. Yan, L. Xia, L. Xu, L. Deng, and G. Jin. A comparative study to determine the association of gut microbiome with schizophrenia in Zhejiang, China. BMC Psychiatry, 22(1):731, Nov 2022.

67. X. Yuan, R. Chen, Y. Zhang, X. Lin, and X. Yang. Sexual dimorphism of gut microbiota at different pubertal status. Microb Cell Fact, 19(1):152, Jul 2020.

68. N. Segata, J. Izard, L. Waldron, D. Gevers, L. Miropolsky, W. S. Garrett, and C. Huttenhower. Metagenomic biomarker discovery and explanation. Genome Biol, 12(6):R60, Jun 2011.

69. C. Yang, D. Mills, K. Mathee, Y. Wang, K. Jayachandran, M. Sikaroodi, P. Gillevet, J. Entry, and G. Narasimhan. An eco-informatics tool for microbial community studies: supervised classification of Amplicon Length Heterogeneity (ALH) profiles of 16S rRNA. J Microbiol Methods, 65(1):49–62, Apr 2006.

70. Pourhoseingholi Mohamad Amin, Baghestani Ahmad Reza, and Vahedi Mohsen. How to control confounding effects by statistical analysis. 2012.

71. F. S. Nahm. Receiver operating characteristic curve: overview and practical use for clinicians. Korean J Anesthesiol, 75(1):25–36, Feb 2022.

72. Corinna Cortes and Mehryar Mohri. Confidence intervals for the area under the roc curve. In L. Saul, Y. Weiss, and L. Bottou, editors, Advances in Neural Information Processing Systems, volume 17. MIT Press, 2004.

73. K. lu and G. Aksel. Receiver operating characteristic curve analysis in diagnostic accuracy studies: A guide to interpreting the area under the curve value. Turk J Emerg Med, 23(4):195–198, 2023.

74. Elizabeth R. DeLong, David M. DeLong, and Daniel L. Clarke-Pearson. Comparing the areas under two or more correlated receiver operating characteristic curves: a nonparametric approach. Biometrics, 44 3:837–45, 1988.

75. Curtis Huttenhower, Dirk Gevers, Rob Knight, Sahar Abubucker, Jonathan Badger, Asif Chinwalla, Heather Huot Creasy, Earl AM, Michael Fitzgerald, Robert Fulton, Michelle Giglio, Kymberlie Pepin, Lobos EA, Ramana Madupu, Vincent Magrini, John Martin, Makedonka Mitreva, Muzny DM, Sodergren EJ, and Owen White. The human microbiome project (hmp) consortium. structure, function and diversity of the healthy human microbiome. nature 486: 207-214. Nature, 486:207–214, 06 2012.

76. J. G. Natalini, S. Singh, and L. N. Segal. The dynamic lung microbiome in health and disease. Nat Rev Microbiol, 21(4):222–235, Apr 2023.

77. Ayako Muraoka, Miho Suzuki, Tomonari Hamaguchi, Shinya Watanabe, Kenta Iijima, Yoshiteru Murofushi, Keiko Shinjo, Satoko Osuka, Yumi Hariyama, Mikako Ito, Kinji Ohno, Tohru Kiyono, Satoru Kyo, Akira Iwase, Fumitaka Kikkawa, Hiroaki Kajiyama, and Yutaka Kondo. Fusobacterium infection facilitates the development of endometriosis through the phenotypic transition of endometrial fibroblasts. Science translational medicine, 15:eadd1531, 06 2023.

78. M. Labetoulle, C. Baudouin, J. M. Benitez Del Castillo, M. Rolando, M. Rescigno, E. M. Messmer, and P. Aragona. How gut microbiota may impact ocular surface homeostasis and related disorders. Prog Retin Eye Res, 100:101250, May 2024.

79. S. Shivaji. A systematic review of gut microbiome and ocular inflammatory diseases: Are they associated? Indian J Ophthalmol, 69(3):535–542, Mar 2021.

80. Jerome Ozkan and Mark Willcox. The ocular microbiome: Molecular characterization of a unique and low microbial environment. Current Eye Research, 44, 01 2019.

81. Jose Alvaro P Gomes, Luciana Frizon, and Vanessa F Demeda. Ocular surface microbiome in health and disease. The Asia-Pacific Journal of Ophthalmology, 9(6):505–511, 2020.

82. Mariona Pinart, Andreas Dötsch, Kristina Schlicht, Matthias Laudes, Jildau Bouwman, Sofia K Forslund, Tobias Pischon, and Katharina Nimptsch. Gut microbiome composition in obese and non-obese persons: a systematic review and meta-analysis. Nutrients, 14(1):12, 2021.

83. Pasquale Aragona, Christophe Baudouin, Jose M Benitez Del Castillo, Elisabeth Messmer, Stefano Barabino, Jesus Merayo-Lloves, Francoise Brignole-Baudouin, Leandro Inferrera, Maurizio Rolando, Rita Mencucci, et al. The ocular microbiome and microbiota and their effects on ocular surface pathophysiology and disorders. Survey of ophthalmology, 66(6):907–925, 2021.

84. Xiaofeng Wen, Li Miao, Yuhua Deng, Paul W Bible, Xiao Hu, Yanli Zou, Yu Liu, Shixin Guo, Juanran Liang, Tingting Chen, et al. The influence of age and sex on ocular surface microbiota in healthy adults. Investigative ophthalmology & visual science, 58(14):6030–6037, 2017.

85. J. G. Kers and E. Saccenti. The Power of Microbiome Studies: Some Considerations on Which Alpha and Beta Metrics to Use and How to Report Results. Front Microbiol, 12:796025, 2021.

86. B. Fritz, A. Jenner, S. Wahl, C. Lappe, A. Zehender, C. Horn, F. Blessing, M. Kohl, F. Ziemssen, and M. Egert. A view to a kill? - Ambient bacterial load of frames and lenses of spectacles and evaluation of different cleaning methods. PLoS One, 13(11):e0207238, 2018.

87. H. Dziewiecka, H. S. Buttar, A. Kasperska, J. Ostapiuk-Karolczuk, M. Domagalska, J. ń, and A. ska Stejnborn. Physical activity induced alterations of gut microbiota in humans: a systematic review. BMC Sports Sci Med Rehabil, 14(1):122, Jul 2022.

88. J. Philip Karl, Lee M. Margolis, Elisabeth H. Madslien, Nancy E. Murphy, John W. Castellani, Yngvar Gundersen, Allison V. Hoke, Michael W. Levangie, Raina Kumar, Nabarun Chakraborty, Aarti Gautam, Rasha Hammamieh, Svein Martini, Scott J. Montain, and Stefan M. Pasiakos. Changes in intestinal microbiota composition and metabolism coincide with increased intestinal permeability in young adults under prolonged physiological stress. American Journal of Physiology-Gastrointestinal and Liver Physiology, 312(6):G559–G571, 2017. PMID: 28336545.

89. Ruwen Zhou, Siu Kin Ng, Joseph Jao Yiu Sung, Wilson Wen Bin Goh, and Sunny Hei Wong. Data pre-processing for analyzing microbiome data – a mini review. Computational and Structural Biotechnology Journal, 21:4804–4815, 2023.

